# SARS-CoV-2 screening in patients in need of urgent inpatient treatment in the Emergency Department (ED) by digitally integrated point-of-care PCR: A clinical cohort study

**DOI:** 10.1101/2022.01.01.22268603

**Authors:** Martin Möckel, Myrto Bolanaki, Jörg Hofmann, Angela Stein, Jennifer Hitzek, Fabian Holert, Antje Fischer-Rosinský, Anna Slagman

**Affiliations:** Department of Emergency and Acute Medicine, Charité – Universitätsmedizin Berlin, Campus Mitte and Virchow; Institute of Virology, Charité - Universitätsmedizin Berlin and Labor Berlin - Charité Vivantes GmbH

## Abstract

Patients in need of urgent inpatient treatment were recruited prospectively. A rapid point of care PCR test (POC-PCR; Liat®) for SARS-CoV2 was conducted in the ED and a second PCR-test from the same swab was ordered in the central laboratory (CL-PCR). POC-PCR analyzers were digitally integrated in the laboratory information system.

Overall, 160 ED patients were included. A valid POC-PCR-test result was available in 96.3% (n=154) of patients. N=16 patients tested positive for SARS-CoV-2 (10.0%). The POC PCR test results were available within 102 minutes (median, IQR: 56-211), which was significantly earlier compared to the CL PCR (811 minutes; IQR: 533-1289, p < 0.001). The diagnostic accuracy of the POC-PCR test was 100%. The implementation and digital LIS integration was successfully done. Staff satisfaction with the POC process was high.

The POC-PCR testing in the emergency department is feasible and shows a very high diagnostic performance.

Trial registration: DRKS00019207

## Introduction

The COVID-19 pandemic put major challenges on Emergency Departments (ED) worldwide(1). One major aspect was that all urgent patients had to be treated mostly without knowing the SARS-CoV-2 status as laboratory based polymerase chain reaction (PCR)-test usually took at least 6-8 hours of turn-around-time (TAT)(2). The introduction of rapid antigen testing had the advantage of faster results on the cost of false negative results. Although some authors thought that false negative rapid antigen tests may due to low virus concentrations only(3), we could show that this is not true in clinical practice and that among false negatives are infectious patients with high virus load(4). Thus, specifically for emergencies, which require immediate intervention or operation and probably airway-management like patients with multiple trauma, acute myocardial infarction or stroke, a rapid, point-of-care (POC) SARS-CoV-2 PCR test would be of utmost importance(5).

POC testing is standard in the ED for clinical chemistry parameters like lactate, electrolytes or cardiac troponin. For molecular diagnostics there have been first attempts with influenza POC testing(6). Due to the high importance of the test results, which touch regulatory aspects like hygiene rules and due the fast disposition and transfers (i.e. operating theatre, ICU) of the high urgent patients, a digital integration of the POC device in the LIS seems mandatory. In addition, strict testing rules to guarantee correct results, staff safety and reliable results information for all involved health care professionals have to be applied. Finally, the used instrument and assay need to provide reliable results under routine conditions(7).

Therefore, in the current study, a SARS-CoV-2 PCR POC analyzer was digitally integrated in the LIS and tests were run by ED staff in high urgent patients.

## Methods

### Patient population

All inpatients are screened for SARS-CoV-2 in our ED. Patients were prospectively recruited once it became clear that an acute intervention of any kind was indicated.

### Data collection and endpoints

Clinical characteristics and in-hospital follow-up information of all included patients were extracted from electronic medical records.

The primary endpoint of the analyses was turn-around-time (TAT) of the POC-PCR as compared to TAT of the CL-PCR in relation to the first intervention. Secondary endpoints were diagnostic performance, staff satisfaction with test handling and transfer of test results, patient safety and description of further clinical endpoints (e.g. stay on the intensive care unit (ICU), extra-corporal membrane oxygenation (ECMO), in-hospital mortality).

### Statistical Analysis

Descriptive analyses included the calculation of relative and absolute frequencies as well as median and interquartile range (IQR). Statistical differences were calculated using the Chi-square test for categorical variables and the Mann-Whitney test for continuous variables. A p-value of less than 0.05 was considered statistically significant. Due to the exploratory nature of the analysis, no corrections were made for multiple testing.

### Digital integration of POC analyzers

Liat®POC analyzers (Roche Molecular Systems) were connected to the laboratory information system via the hospital LAN system and Roche infinity POC middleware. The POC test was ordered via the laboratory channel, a barcode was generated and put on the test tube. A triple test cartridge (SARS-CoV-2, influenza A & B) was used. The barcode was scanned by the POC instrument and identified the order. Once the analyzer completed the test run, results were automatically transferred. In the central (molecular biology) laboratory the POC test results were automatically confirmed and transferred to the hospital information system. The LIS-HIS interface was refreshed every 15min. Thus with a 20min testing time on the Liat® analyzers, the SARS-CoV-2 POC test results were digitally available hospital wide within a maximum of 35min after the start of test.

### Hygiene concept and test execution

As the work with potential contagious samples require specific hygiene rules, these were set up according to national regulations. One challenge was that these rules allow the SARS-CoV-2 sample handling outside a laboratory setting only, if it is performed by the protected person who did the swab. Therefore, we set up the procedure in the way that the POC test cartridge was filled in the isolation room of the patient. Once the cartridge was closed, it could be handled by another member of the staff, who started the test run of the instrument at a central location of the ED. The details of the test procedure are available in the supplemental material (S1). For standardization purposes known virus preparations and dilutions were measured. The transport media was directly pipetted into the test cartridge without further dilution in a buffer.

The digitally integrated use of the POC instrument in the ED is summarized in the figure 1.

**Figure 1.**
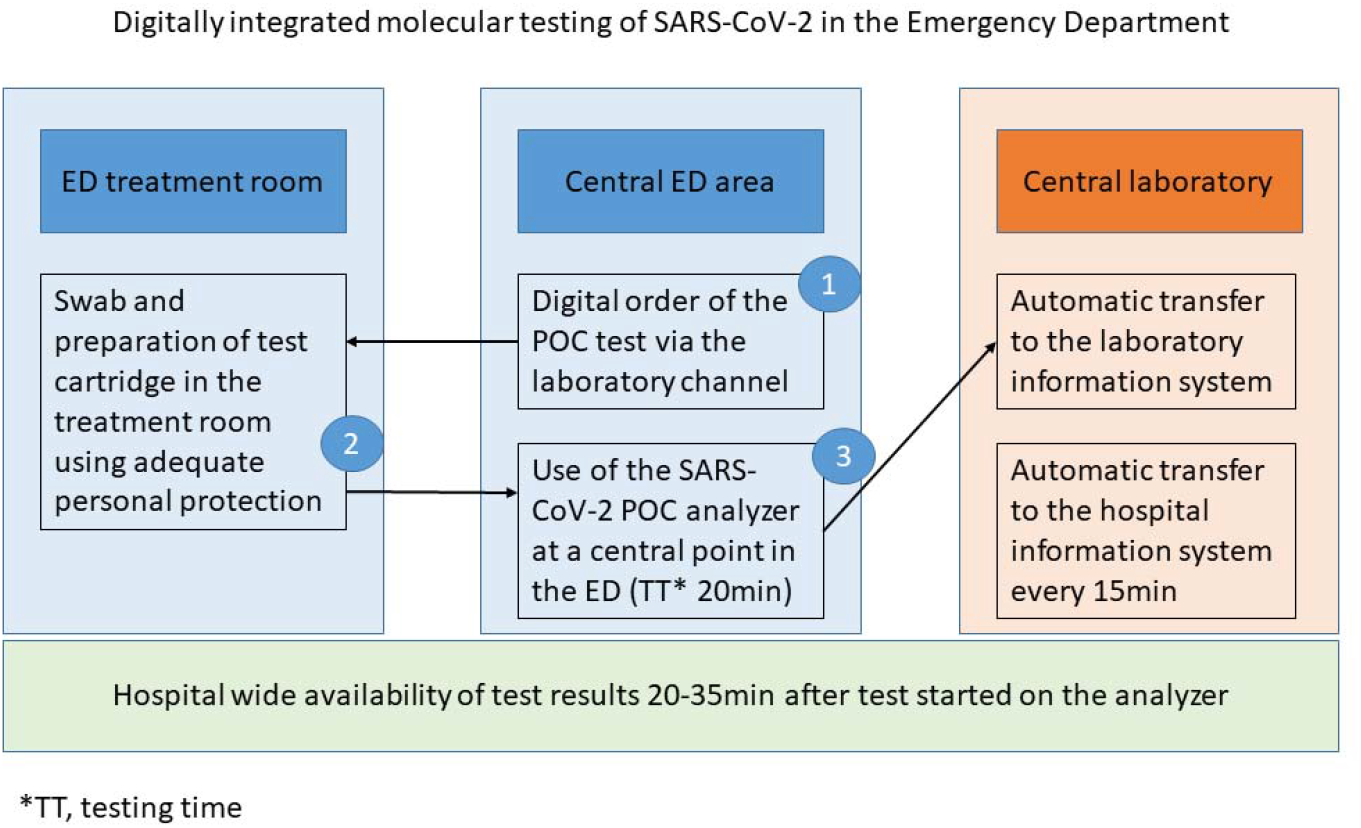
Process of the digitally integrated SARS-CoV-2 POC.

### Ethics

The Ethics committee of the Charité approved the study as an amendment to a previous POC influenza investigation (EA2/204/19). The study was registered in the German Clinical Trials Registry (DRKS00019207).

## Results

### Patient characteristics

The population of patients in urgent need of inpatient treatment (n=160) consisted of 10.0% of patients tested positive for SARS-CoV-2 in POC-PCR-testing (n=16) and 86.3% who were tested negative (86.3%). In 6 patients an invalid result occurred. Figure 2 shows the patient flow diagram.

**Figure 2.**
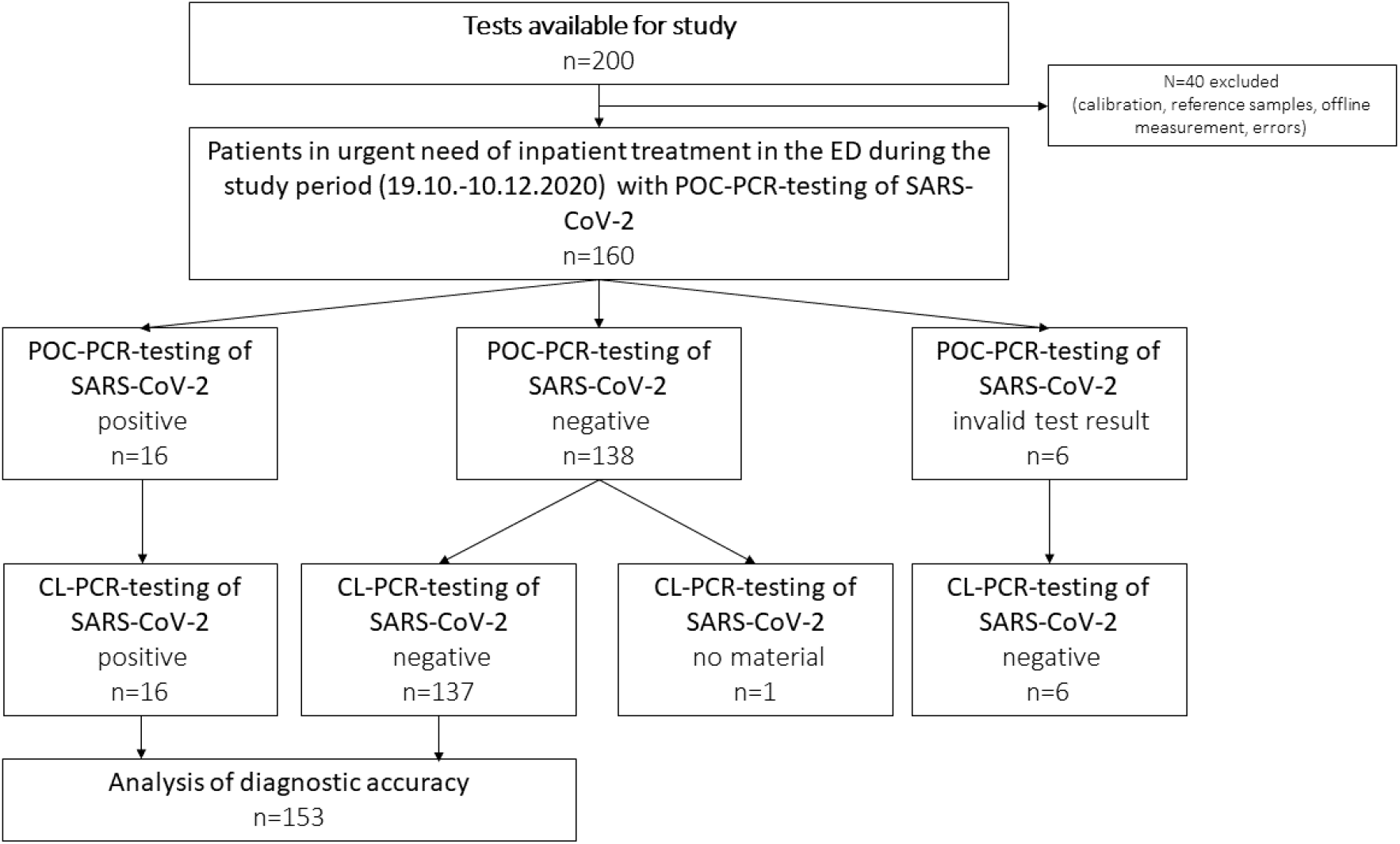
Patient flow diagram. For this implementation and feasibility study, n=200 tests were available.

Of all patient, 43.1% (n=69) were women and the median age was 68 years (IQR: 51-78).

In confirmed SARS-CoV-2 cases by POC-PCR-testing, the proportion of female patients was 50.0% (n=8) as compared to 41.3% (n=57) in patients who were tested negative for SARS-CoV-2 (table 1a). The most common symptoms in SARS-CoV-2 positive cases were Dyspnea (37.5%; n=6), fever and cough (25.0%; n=7 each) table 1b. Laboratory parameters are depicted in table 1b. Diagnostic accuracy of POC-PCR was compared to CL-PCR (reference method) in all patients in urgent need of inpatient treatment. The resulting diagnostic performance values are: sensitivity 100% (95%-CI: 79-100), specificity 100% (95%-CI: 97-100), PPV and NPV 100% (table 2). The most common reasons for urgent need of inpatient treatment were admission to ICU (25.0%; n=40), urgent operation (22.5%, n=36) and stroke (18.1%; n=29; table 3).

**Table 1.**
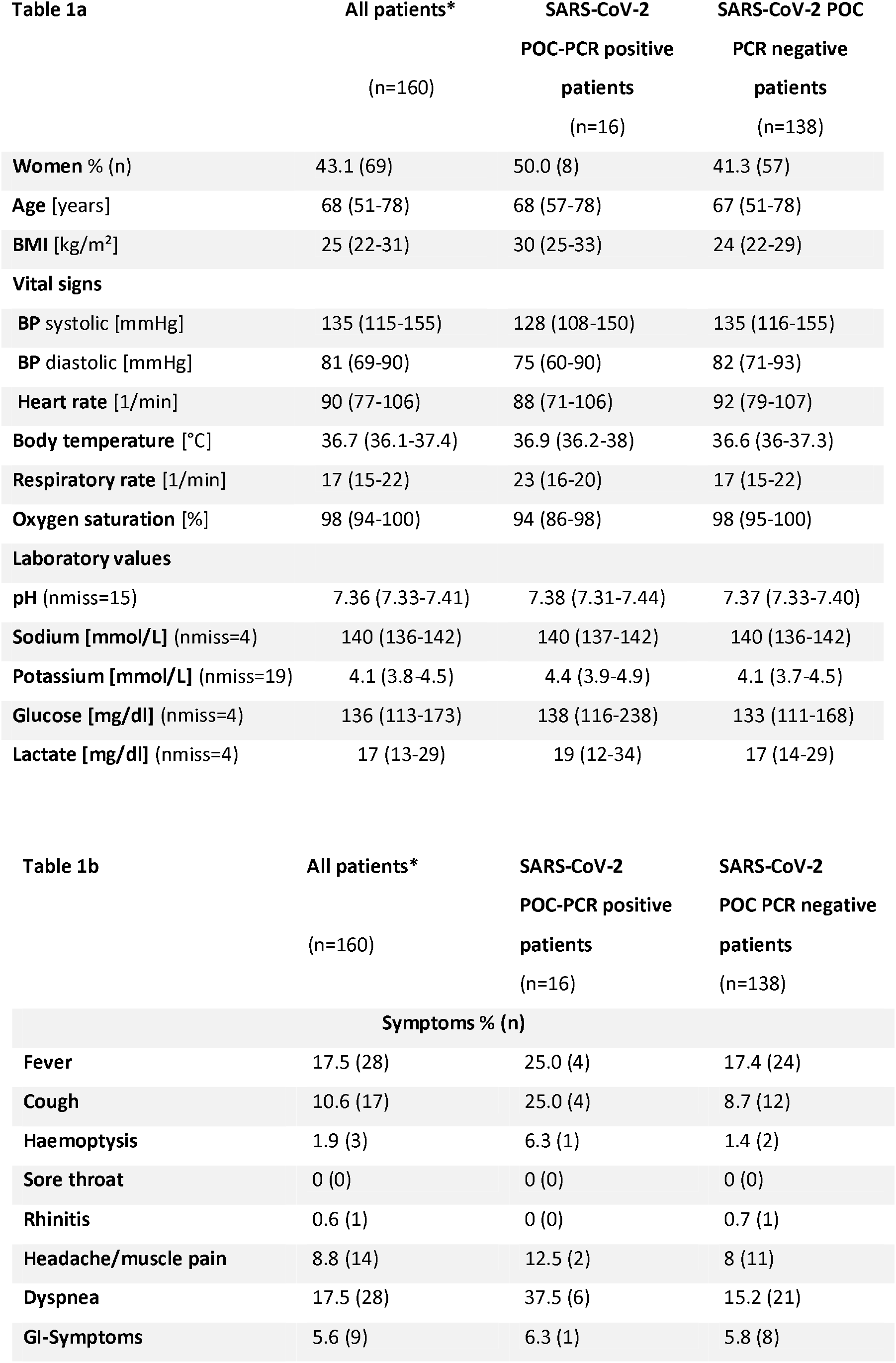
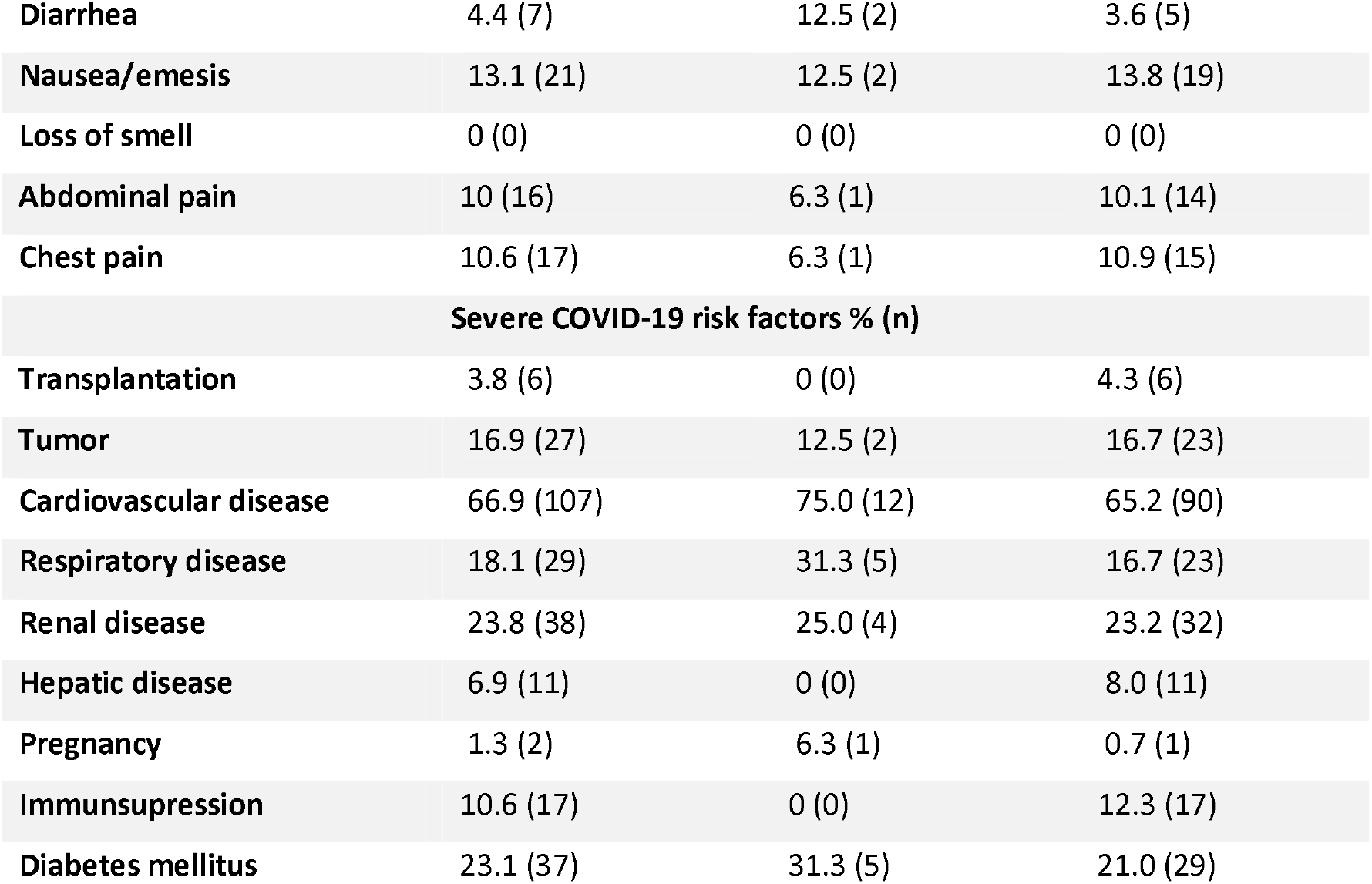
a. Demographic and clinical characteristics of all patients who were tested positive or negative for SARS-CoV-2 by POC-PCR; values are median (25%-75% percentiles); *, in n=6 patient an invalid result occurred on the POC instrument. BMI, body mass index; BP, systemic arterial blood pressure. b. Symptoms and risk factors of all participants and patients who were tested positive or negative for SARS-CoV-2 by POC-PCR; *, in n=6 patient an invalid result occurred on the POC instrument.

| **Table 1a** | **All patients\*** | **SARS-CoV-2** | **SARS-CoV-2 POC** |
| --- | --- | --- | --- |
|  |  | **POC-PCR positive** | **PCR negative** |
|  | (n=160) | **patients** | **patients** |
|  |  | (n=16) | (n=138) |
| **Women % (n)** | 43.1 (69) | 50.0 (8) | 41.3 (57) |
| **Age [years]** | 68 (51-78) | 68 (57-78) | 67 (51-78) |
| **BMI [kg/m<sup>2</sup>]** | 25 (22-31) | 30 (25-33) | 24 (22-29) |
| **Vital signs** |  |  |  |
| **BP systolic [mmHg]** | 135 (115-155) | 128 (108-150) | 135 (116-155) |
| **BP diastolic [mmHg]** | 81 (69-90) | 75 (60-90) | 82 (71-93) |
| **Heart rate [1/min]** | 90 (77-106) | 88 (71-106) | 92 (79-107) |
| **Body temperature [°C]** | 36.7 (36.1-37.4) | 36.9 (36.2-38) | 36.6 (36-37.3) |
| **Respiratory rate [1/min]** | 17 (15-22) | 23 (16-20) | 17 (15-22) |
| **Oxygen saturation [%]** | 98 (94-100) | 94 (86-98) | 98 (95-100) |
| **Laboratory values** |  |  |  |
| **pH (nmiss=15)** | 7.36 (7.33-7.41) | 7.38 (7.31-7.44) | 7.37 (7.33-7.40) |
| **Sodium [mmol/L] (nmiss=4)** | 140 (136-142) | 140 (137-142) | 140 (136-142) |
| **Potassium [mmol/L] (nmiss=19)** | 4.1 (3.8-4.5) | 4.4 (3.9-4.9) | 4.1 (3.7-4.5) |
| **Glucose [mg/dl] (nmiss=4)** | 136 (113-173) | 138 (116-238) | 133 (111-168) |
| **Lactate [mg/dl] (nmiss=4)** | 17 (13-29) | 19 (12-34) | 17 (14-29) |

| **Table 1b** | **All patients\*** | **SARS-CoV-2** | **SARS-CoV-2** |
| --- | --- | --- | --- |
|  |  | **POC-PCR positive** | **POC PCR negative** |
|  | (n=160) | **patients** | **patients** |
|  |  | (n=16) | (n=138) |
| **Symptoms % (n)** |  |  |  |
| **Fever** | 17.5 (28) | 25.0 (4) | 17.4 (24) |
| **Cough** | 10.6 (17) | 25.0 (4) | 8.7 (12) |
| **Haemoptysis** | 1.9 (3) | 6.3 (1) | 1.4 (2) |
| **Sore throat** | 0 (0) | 0 (0) | 0 (0) |
| **Rhinitis** | 0.6 (1) | 0 (0) | 0.7 (1) |
| **Headache/muscle pain** | 8.8 (14) | 12.5 (2) | 8 (11) |
| **Dyspnea** | 17.5 (28) | 37.5 (6) | 15.2 (21) |
| **GI-Symptoms** | 5.6 (9) | 6.3 (1) | 5.8 (8) |
| Diarrhea | 4.4 (7) | 12.5 (2) | 3.6 (5) |
| Nausea/emesis | 13.1 (21) | 12.5 (2) | 13.8 (19) |
| Loss of smell | 0 (0) | 0 (0) | 0 (0) |
| Abdominal pain | 10 (16) | 6.3 (1) | 10.1 (14) |
| Chest pain | 10.6 (17) | 6.3 (1) | 10.9 (15) |
| **Severe COVID-19 risk factors % (n)** |  |  |  |
| Transplantation | 3.8 (6) | 0 (0) | 4.3 (6) |
| Tumor | 16.9 (27) | 12.5 (2) | 16.7 (23) |
| Cardiovascular disease | 66.9 (107) | 75.0 (12) | 65.2 (90) |
| Respiratory disease | 18.1 (29) | 31.3 (5) | 16.7 (23) |
| Renal disease | 23.8 (38) | 25.0 (4) | 23.2 (32) |
| Hepatic disease | 6.9 (11) | 0 (0) | 8.0 (11) |
| Pregnancy | 1.3 (2) | 6.3 (1) | 0.7 (1) |
| Immunosuppression | 10.6 (17) | 0 (0) | 12.3 (17) |
| Diabetes mellitus | 23.1 (37) | 31.3 (5) | 21.0 (29) |

**Table 2.** Diagnostic accuracy of POC-PCR as compared to CL-PCR (reference method) in patients in urgent need for treatment. The resulting diagnostic performance values are: sensitivity 100% (95%-CI: 79-100), specificity 100% (95%-CI: 97-100), PPV and NPV 100%.

| Table 2 | SARS-CoV-2 CL positive | Border-line | Weak | Inter-mediate | High | SARS-CoV-2 CL negative | No material | Total |
| --- | --- | --- | --- | --- | --- | --- | --- | --- |
| SARS-CoV-2 POC positive | 16 |  |  |  |  | 0 |  | 16 |
| Borderline |  | 2 | 1 | 0 | 0 |  |  |  |
| Weak |  | 0 | 4 | 1 | 1 |  |  |  |
| Intermediate |  | 0 | 0 | 0 | 2 |  |  |  |
| High |  | 0 | 0 | 0 | 5 |  |  |  |
| SARS-CoV-2 POC negative | 0 |  |  |  |  | 137 | 1 | 138 |
| Error |  |  |  |  |  | 6 |  | 6 |
| Total | 16 |  |  |  |  | 143 | 1 | 160 |

**Table 3.** Clinical endpoints of all patients who were tested positive or negative for SARS-CoV-2 by POC-PCR; *, in n=6 patient an invalid result occurred on the POC instrument.

| Table 3 | All patients<br>(n=160) | SARS-CoV-2 POC-PCR positive<br>patients<br>(n=16) | SARS-CoV-2 POC-PCR<br>negative<br>patients<br>(n=138) |
| --- | --- | --- | --- |
| **Inclusion criteria for inpatient treatment % (n)** |  |  |  |
| Acute severe trauma | 5.6 (9) | 6.3 (1) | 5.8 (8) |
| Stroke | 18.1 (29) | 12.5 (2) | 18.1 (25) |
| Myocardial Infarction | 8.8 (14) | 0 (0) | 10.1 (14) |
| Urgent operation | 22.5 (36) | 0 (0) | 25.4 (35) |
| Intervention needs<br>within 6 hours after<br>presentation | 6.9 (11) | 0 (0) | 8.0 (11) |
| ICU admission needed<br>within 6 hours after<br>presentation | 25.0 (40) | 62.5 (10) | 20.3 (28) |
| Other reasons | 13.1 (21) | 18.8 (3) | 12.3 (17) |
| **Clinical Course** |  |  |  |
| Intensive Care Unit % (n) | 48.8 (78) | 50.0 (8) | 46.4 (64) |
| Intubation % (n) | 15.6 (25) | 18.8 (3) | 13.8 (19) |
| Death % (n)* | 10 (16) | 25 (4) | 7.2 (10) |

### Primary endpoint

The POC PCR test was available within 102 minutes (median, IQR: 56-211) after admission, which was significantly earlier compared to the CL PCR (811 min; IQR: 533-1289, p < 0.001). In 77.4% of patients the first relevant intervention depending on the inclusion criteria (acute severe trauma, stroke, myocardial infarction, operation, intervention needed within 6 hours after presentation, ICU admission needed within 6 hours after presentation) was done within 6 hours (n=113). The POC test result was available before the intervention in 92.1% (n=129) compared to 5.4% test results from the CL (n=8). There were no positive influenza tests.

### Standard sample measurements

We measured standard samples provided by the institute of virology with the following results.

#### Ct-values of the calibration samples (series on the same Liat® instrument)

G21124.1 (GE/mL 1.93E+07) = 18.6 [Ct ref. Cobas 6800: 25.2]

G21125.1 (GE/mL 2.76E+06) = 21.1 [Ct ref. Cobas 6800: 28.2]

G21126.1 (GE/mL 2.43E+05) = 24.5 [Ct ref. Cobas 6800: 32.0]

#### Ct-values of the dilutions

G21126-2 (1:10) = Ct 28.0

G21126-2 (1:100) = Ct 31.5

G21126-2 (1:1,000) Ct 35.5

G21126-2 (1:10,000) Ct 42 = negative

### Diagnostic performance

Ct-values were converted into estimated virus concentrations per mL buffer using the standards listed above. Levels below 10^6^ per mL buffer are considered as “weak positive”, between 10^6^ and 10^7^ are considered as “intermediate” and over 10^7^ as high. Calculated virus concentrations below 10^4^ are labelled as “borderline” (repeat measurement after 24-48h).

### Clinical endpoints

Clinical endpoint are listed in table 3.

### Staff satisfaction questionnaire

The questionnaire comprised of 11 questions and the opportunity of free text. The questionnaire is available in the supplement 2 (S2). N=37 members of staff, who were involved in the operation of the POC instrument participated. 89% of respondents performed more than 10 measurements with the Liat in and outside of the current study. 73% performed more than 25 measurements and stated that they can integrate the POC testing very well into the treatment routine in the ED. More than 70% of respondents stated they were satisfied with the usability of the devices and 62% were satisfied with sample handling. In contrast 5% were unsatisfied with sample handling: holding the cartridges and pipettes simultaneously as well as the need to pipette beside the patient were points of criticism.

In this current context 46% of the respondents were unsatisfied with hygienic conditions of sample processing. Main aspects for concerns were the location of the devices and the safety for employees and patients. 57% of respondents stated they were satisfied with display of results, 38% were undecided. Medical employees indicated that there should be greater ability to print the results directly at the POC instrument. Furthermore the results of POC testing had various effects. For one, they had an impact on interactions with the patients for 81% of the respondents, in addition, 86% of the respondents stated that the available results were valued by further departments/wards.

## Discussion

The current study confirms(8, 9) the high accuracy of the Liat® SARS-CoV-2 POC PCR test and for the first time the feasibility of use in a large and busy ED with digital integration in the laboratory and hospital information systems (see figure 1).

### POC test performance

The use of POC systems are always intended to shorten the turn-around-time (TAT). As in other studies, this could be shown here. Nevertheless, more important than the pure TAT is (1) the “time to actionable result” time of the attending physicians and (2) whether any important clinical action follows a POC test result. In our study we could clearly show that a majority of SARS-CoV-2 POC test results was available before an urgent care process has been started. Nevertheless, although the pure testing time of the instrument is 20min, the median time from admission to result was 102min, reflecting that there has been a delay in the implementation of the ED test strategy at the time of this study. As the test strategy is now standard in our institution and test capacities are sufficient, this time may be shorter in current routine care. The accuracy of the test results was 100%, although semi-quantification revealed some differences (see table 2). The sensitivity of the POC instrument seems to be comparable to the reference standard, which is an advantage compared to other POC PCR systems, where 1-2% false negatives are reported(7).

### Digital integration

Digital integration is a success factor under several aspects: (1) The digital order of the test including a standard label for the test tube ensures that the risk to mix up samples is minimized. (2) As the major incentive for the SARS-CoV-2 POC PCR is to speed up a complex pathway for emergency patients, it is mandatory that the test results are available for treating units including the operating theatre, intensive care and stroke units etc. In addition, tests need to be repeated and for this, it is most beneficial to have the results of the initial POC testing in the same system as the other laboratory test results. (3) The laboratory with experts for molecular test are automatically in the loop and contribute to quality management. Meanwhile we transfer also the cycle threshold values and a standard laboratory report is done on the basis of the POC test. (4) The POC PCR is a more expensive measure and reimbursement needs to be complete. Having all results in the central system ensures that no test is overseen. We believe that the complete digital integration should be the standard of POC in the ED in general and have established this way of collaboration with the central laboratory earlier for POC of Troponin T(10).

### Staff satisfaction

The use of high end POC devices in the ED requires trained staff, who are able to follow demanding test protocols as in this study due to hygiene rules as described in detail above (see also S1 for details). Thus, it is most important to assess the staff perspective on innovative POC test strategies. The staff was mostly satisfied with the technical aspects including sample handling and the ease of use of the instrument. A major concern, expressed by 46% of the participants were hygiene aspects of the test procedure. Although the test process follows strictly the regulations, many nurses feel uneasy with the step of pipetting a potential infectious sample although being sufficiently protected. We think that one solution of for this challenge could be the use of inactivating lysis buffer in the test tubes(11). Currently, only transport media and saline are approved for the use with the instrument.

Finally, the majority of participants valued that all admission processes and the interaction with patients and colleagues is supported by the POC PCR.

### Perspectives

The following future developments are necessary for the continuation of molecular POC testing in the ED.

1. Use of tubes with inactivating lysis buffer, to make the test safer for the users. Alternatively, laboratory workbenches needs to be integrated in the ED, which may be suitable some but no everywhere.
2. Transfer of cycle threshold (Ct) values to the LIS. Meanwhile, the Ct values are used to estimate virus concentrations in the buffer of swabs on the basis of measurement of virus standards. On the peak of the SARS-CoV-2 waves with many positive samples, this has less importance than in lower incidence times, because now, frequently weak positive signals occur and those patients must not be sent to a COVID-19 ward, where they are placed at risk for an infection, which they might not have at admission. This innovation has been actually released and is tested currently in our ED. Ct-values are automatically translated into a medical report of positive test results as being “to be controlled”, “weak positive” [< 10^6^ virus copies/mL buffer] or “strong positive” [> 10^7^ virus copies/mL buffer] or intermediate positive in between of the other two categories.
3. Future epidemiology of SARS-CoV-2 and other virus diseases will require a more differentiated test approach. Thus, in an actual very low incidence phase and no influenza at all, we would prefer to use a single SARS-CoV-2 test. In pediatrics the combination of SARS-CoV-2 and RSV would be interesting and at winter time the current triple test or even the addition of RSV would be beneficial. Finally, for future instrument development, a standardization of the PCR cycles with central laboratory instruments could help.

## Clinical significance

Our study show that a fully digital integrated POC PCR test strategy improves fast and safe emergency processes in urgent patients under pandemic conditions in the ED.

## Conclusions

The POC-PCR testing in the emergency department is feasible and shows a very high diagnostic performance. The digital process integration is key for a high clinical impact in patients with acute indication for inpatient treatment.

## Data Availability

Data are available upon resonable request from

https://versorgungsforschung.charite.de/

## Acknowledgement

We acknowledge Clinical Research Unit (CRU) by Berlin Institute of Health (BIH), 10178 Berlin, Germany for providing support regarding the use of the data management software (RedCap). We thank Dr. Victor Corman, Institute for Virology, Charité – Universitätsmedizin Berlin, providing the standard virus samples.

## Contributorship Statement

All authors were involved in the conception and design of the study. MB, AFR and AS planned the acquisition of data. MB and AS conducted the statistical analysis. JH and ASt set up the connection to the virology laboratory, performed and interpreted the confirmation tests and supervised the study from the laboratory side. FH set up the local process standard and supervised test activities. JHi was responsible for the questionnaire. MM, MB and AS drafted the manuscript and serve as guarantors for the manuscript. All authors were involved in the interpretation of data, critically revised the manuscript for important intellectual content, approved the final version to be published and agreed to be accountable for all aspects of the work. The corresponding author attests that all listed authors meet authorship criteria and that no others meeting the criteria have been omitted.

