## Supplemental material for "SARS-CoV-2 screening in patients in need of urgent inpatient treatment in the Emergency Department (ED) by digitally integrated point-of-care PCR: A clinical cohort study"

### Performing a SARS-CoV-2 / Influenza A+B POC test on the Cobas LIAT

Fabian Holert
November 25, 2020

### COVID-19 in Emergency Medicine Study 5 (COVID-NA 5)

#### Patient inclusion and test order

1. Identification of a suitable patient based on the inclusion criteria.
2. Test order with sender specific profile „POC CoVID (LIAT)“
   → Print of 3 order labels

#### Nasopharyngeal swab and transfer into assay tube

1. Use of a viral sample collection kit, consisting of a cotton swab and viral transport medium
   → Put the first order label on collection tube.

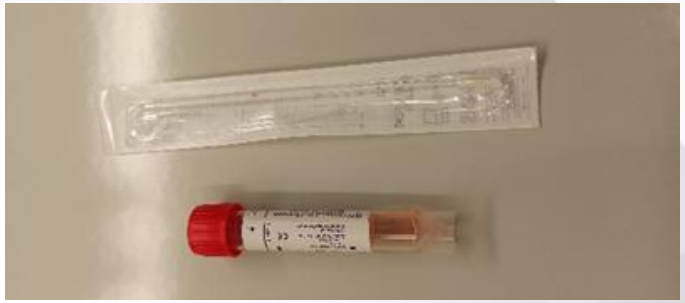

2. Get a SARS-CoV-2 / Influenza A+B test kit from the refrigerator, consisting of an assay tube in a protective sleeve and a single use pipette → Put the second order label on the sleeve.
   *Do not cover the test barcode!*

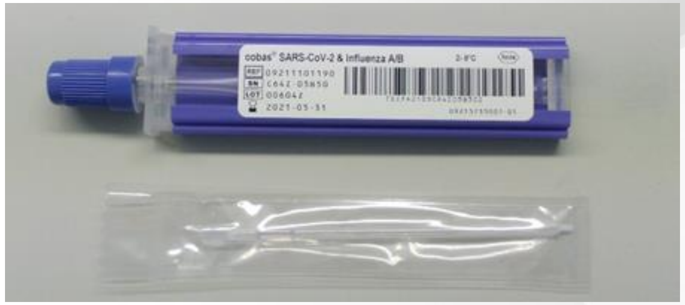

### COVID-19 in Emergency Medicine Study 5 (COVID-NA 5)

<u>For safety reasons, steps 3 + 4 must be performed in the same room:</u>

3. Perform a nasopharyngeal swab according regulation with the necessary safety measures. Transfer the swab into the test tube and let it incubate for at least 1 minute inside the viral transport medium.

4. Place the test tube in a safe stand and open the cap. Open the cap of the assay tube. With the single use pipette, transfer the set amount of transport medium from the test tube into the assay tube (without creating bubbles or perforating the bottom of the assay tube). Dispose of the pipette in a container for infectious waste. Put the caps back onto the test tube and the assay tube.

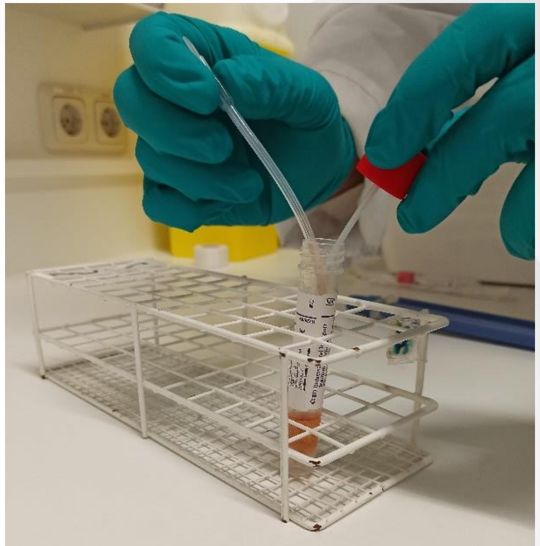

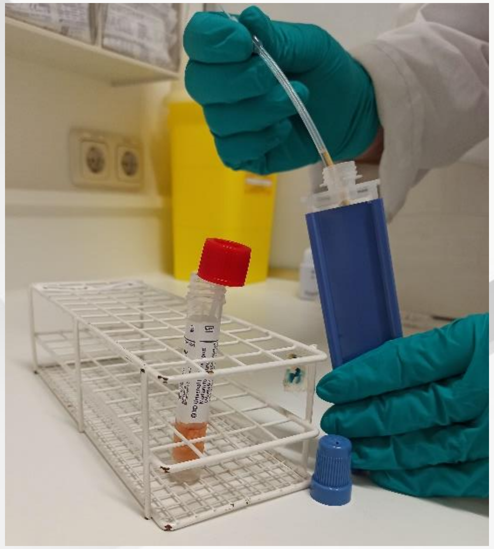

5. Bring the test tube, the assay tube and the third order label to the LIAT analyzer and perform a test run. *Do not hold the filled assay tube upside down.*

#### Perform a run on the LIAT analyzer

1. Put an associated patient label (*not the third order label*) in the result log.

2. Choose “Run Assay” in the main menu.
   If analyzer is logged off, choose “Log on” first and enter name and password.

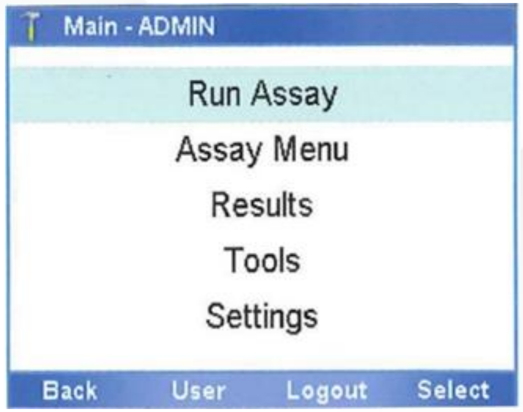


3. Scan the test barcode on the sleeve of the assay tube.

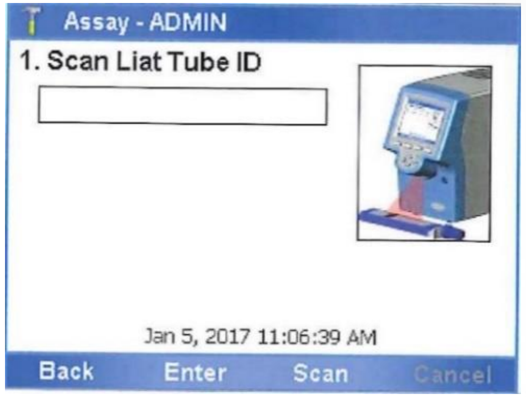


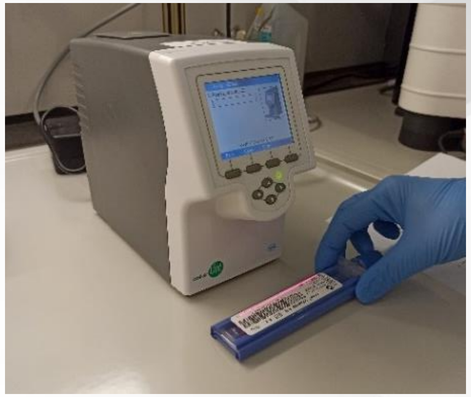

### COVID-19 in Emergency Medicine Study 5 (COVID-NA 5)

4. Scan the order label on the test tube.
   *Compare the order labels on the test tube and on the assay tube, to avoid mix-ups.*

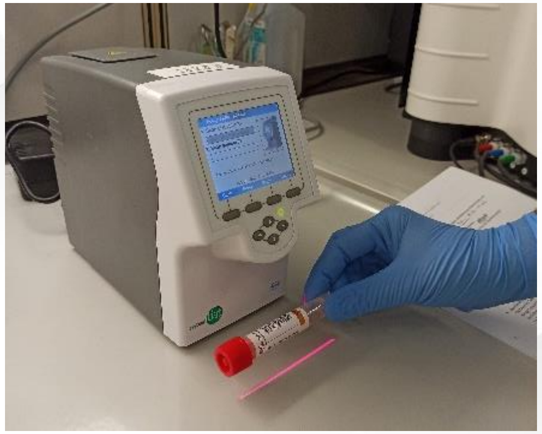

5. Scan the test barcode on the sleeve of the assay tube again.
   The lid on top of the analyzer opens automatically.

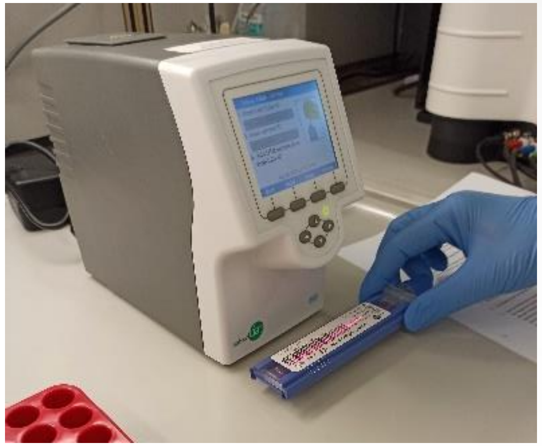

### COVID-19 in Emergency Medicine Study 5 (COVID-NA 5)

6. Remove the assay tube sleeve and insert the assay tube into the analyzer until the tube clicks into place. *This must be done within 20 seconds! It only fits in one way.*

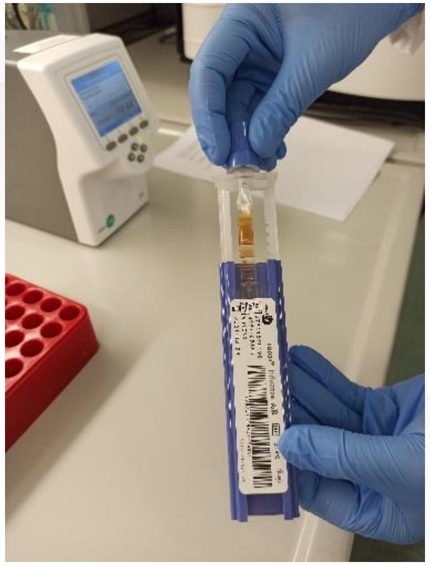
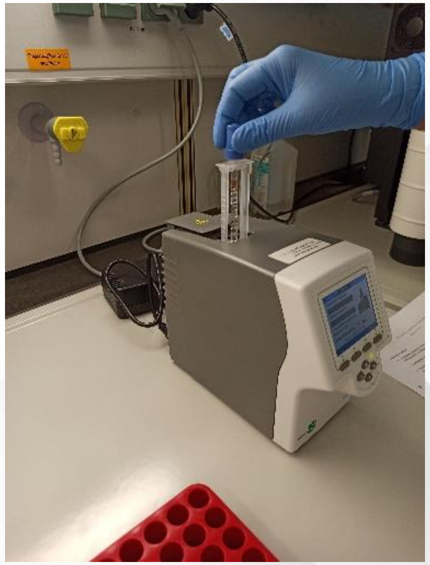
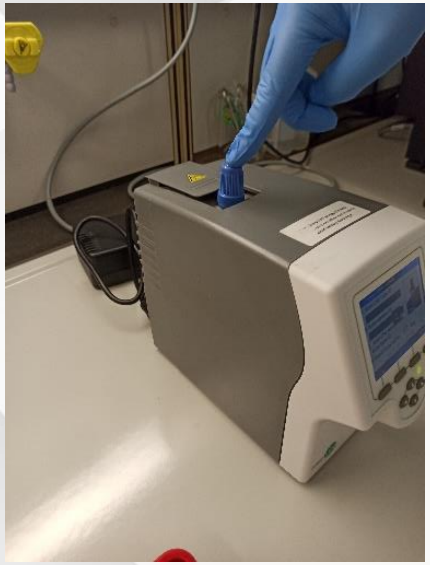

7. The lid closes automatically and processing begins.
The analyzer displays the remaining time.

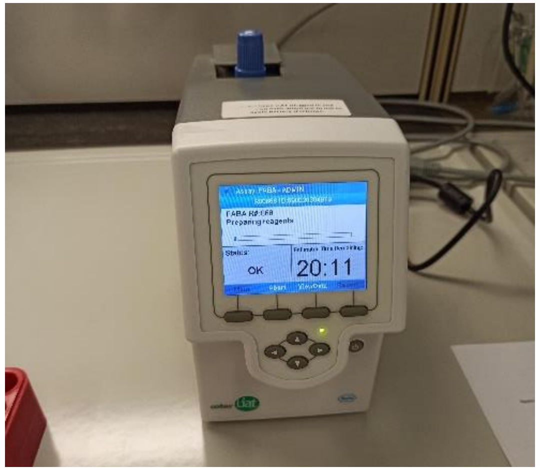

#### Results and sample transport

1. When the assay run is complete, the lid opens and a message asks you to remove the assay tube.

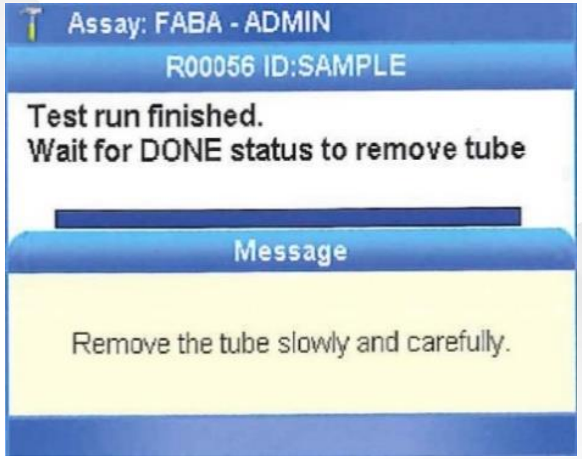


2. Slowly and carefully remove the assay tube and dispose of it in a container for infectious waste.
   The lid closes automatically.

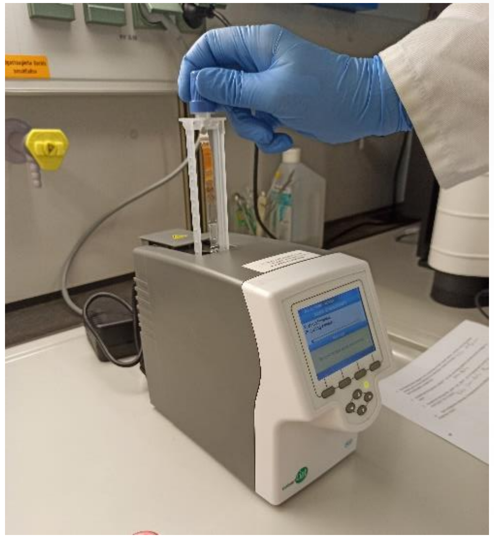

### COVID-19 in Emergency Medicine Study 5 (COVID-NA 5)

3. Choose the “Report” button, to review the results.
   Check the results in the results log.

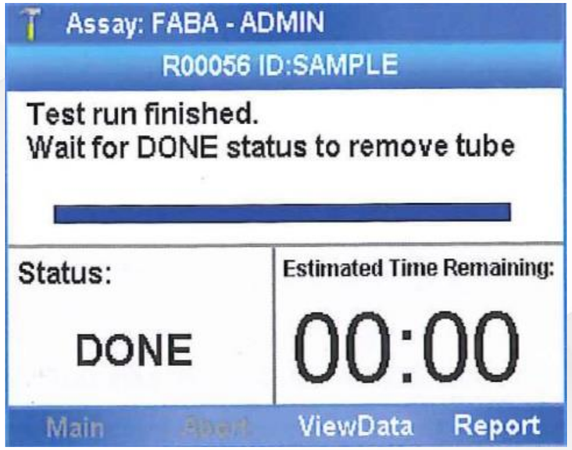


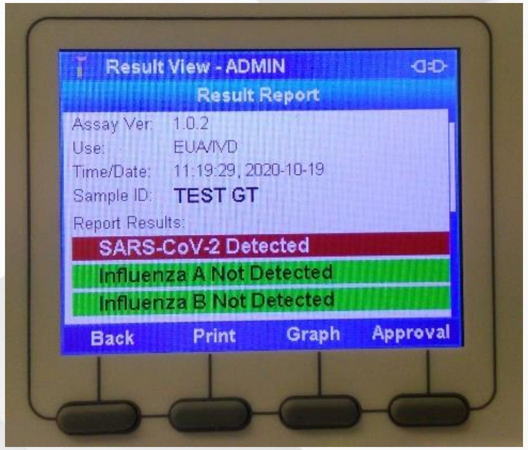


4. Results are sent to a host automatically.

5. Choose the buttons “Back”, then “Main” to get back to the main menu to start a new run.

6. After a successful run, send the test tube to the central laboratory.
